# Epigenetic Mechanisms Regulating the Association between *OR2L13* and Major Psychiatric Disorders

**DOI:** 10.1101/2024.03.04.24303702

**Authors:** Xiujuan Du, Lingli Zhang, Tai Ren, Hua He, Jian Zhao, Fei Li

**Author notes:** Address correspondence to (F.L.); (J.Z.).

## Abstract

**Background:** Previously, population-based cohort studies have identified the association between epigenetic modifications of *OR2L13* related to mental disorders and Gestational diabetes mellitus (GDM). However, the causal nature of these associations remains difficult to establish owing to confounding.

**Aims:** The purpose of the study was to investigate the causal effect of methylation of *OR2L13* and offspring mental health outcomes.

**Method:** We performed two-sample mendelian randomisation to assess the effect of methylation of *OR2L13* on mental disorders. Methylation of 7 CpG sites within *OR2L13* related to GDM from two previous studies were used as exposure. Genome wide significant single nucleotide polymorphisms for methylation of *OR2L13* retrieved from published data were used as instrumental variables. Their causal impact on major psychiatric disorders was assessed using summary-level data mostly from the Psychiatric Genomics Consortium.

**Results:** Lower *OR2L13* methylation was casually associated with a higher risk of PD in offspring [cg03748376: odds ratio (OR)=0.81, 95% confidence interval (CI) =0.68–0.97, P =0.02]. However, little evidence was found for a causal relationship between the methylation levels of *OR2L13* and autism spectrum disorder (ASD), attention deficit/hyperactivity disorder (ADHD), schizophrenia (SCZ), major depressive disorder (MDD), bipolar disorder (BD) and obsessive-compulsive disorder (OCD).

**Conclusions:** Evidence from our study supported a causal effect of lower *OR2L13* methylation on PD risk.

## Introduction

Gestational diabetes mellitus (GDM) is glucose intolerance that begins during pregnancy and associated with adverse neurobehavioral outcomes in offspring, including autism spectrum disorder (ASD), attention deficit/hyperactivity disorder (ADHD) and lower intelligence scores ^1, 2^. In the past decade, growing evidence has shown a link between epigenetic processes, and a range of mental health disorders ^3–5^. Previous population-based cohort studies demonstrated that epigenetic modifications induced by hyperglycemia in utero environment can contribute to adverse health outcomes in offspring ^6–8^. Howe et al. reported that CpG sites of the *OR2L13* promoter (7 probes: cg20433858, cg03748376, cg00785941, cg04028570, cg08260406, cg08944170, cg20507276) were lower hypomethylated in newborns exposed to GDM in utero compared to control subjects in the Pregnancy and Childhood Epigenetics Consortium (including 3,677 mother-neonate pairs from 7 pregnancy cohorts)^6^. The EWAS replication analysis in the Danish National Birth Cohort (DNBC, 9– to 16 – year-old offspring of women with GDM, n=188) further demonstrated that the association can last until at least pre-adolescent or adolescent age. Wong et al. found that one differentially methylated region (DMR) within the *OR2L13* putative gene promoter, contained the ASD-specific differentially methylated probes (DMPs) identified in the case-control analysis of peripheral blood8. However, causal evidence is still limited on the association between methylation of *OR2L13* induced by GDM and offspring mental health outcomes. Previous associations between early life molecular traits and major psychiatric disorders were commonly derived using multivariable regression analysis in observational studies, which is prone to confounding or reverse causation bias ^9^. Mendelian randomisation (MR), a cutting-edge causal inference approach using genetic variants associated with modifiable risk factors as instrumental variables to infer the causal relationship between risk factors and the outcome of interest, has been increasingly used in modern epidemiology research^10, 11^. Thus, in this study, we utilized MR to examine the causal effects of variations in *OR2L13* related methylation levels driven by maternal GDM on a range of major psychiatric disorders.

## Materials and methods

### Exposure

In most of MR studies, maternal genetic variants are used as IVs to test the effect of a pregnancy (intrauterine) exposure on offspring outcomes. Yet, offspring inherit 50% of their genotype from each parent, and offspring genetic variants strongly related to their outcome might be the same as variants used in their mother’s IV. Then the assumption known as InSIDE (instrument strength independent of direct effect) is like to be violated ^12^. Therefore, we used changes of cord blood DNA methylation levels to proxy for adverse intrauterine environment caused by GDM. Based on the findings replicated in independent cohorts mentioned before, we focused on the targeted CpG sites of the *OR2L13* promoter. Genetic variants associated with blood DNA methylation level changes were extracted from the largest DNA methylation-quantitative trait locus (mQTL) analyses, which combined data from 36 population-based cohorts ^13^. This dataset comprises 32,851 European samples, identifying genetic variants associated with DNAm at 420,509 DNAm sites in blood. A total of 248,607 independent cis-mQTL associations (p<1e-8, <1Mb from the DNAm site) were identified in the dataset. Single-nucleotide polymorphisms (SNPs) significantly associated with CpG sites of the *OR2L13* promoter (7 probes) were included in the MR analyses, which can be found in Supplementary Table S1.

### Outcomes

Major psychiatric disorders, including ASD, ADHD, schizophrenia (SCZ), major depressive disorder (MDD), bipolar disorder (BD), obsessive-compulsive disorder (OCD), panic disorder (PD) were examined in this study ^14–20^. Summary-level data of these outcomes were retrieved from several studies (Table 1), among which most were downloaded from the Psychiatric Genomics Consortium (https://pgc.unc.edu/for-researchers/download-results/).

**Table 1.** Characteristics of GWAS summary data on ASD, ADHD, SCZ, MDD, BD, OCD and PD.

| Outcome | First Author | Consortium/cohort | Sample size |
| --- | --- | --- | --- |
| ASD | Matoba et al.<sup>14</sup>,<br>2020 | SPARK & PGC | SPARK, 6222 case-pseudocontrol pairs<br>PGC, Ncase = 18,381 vs.<br>Ncontrol = 27,969 |
| ADHD | Demontis et al.<sup>15</sup>,<br>2019 | PGC | Ncase = 19,099 vs.<br>Ncontrol = 34,194 |
| SCZ | Trubetskoy et al.<sup>16</sup>,<br>2022 | PGC | Ncase = 76,755 vs.<br>Ncontrol = 243,649 |
| MDD | Wray et al.<sup>17</sup>,<br>2018 | PGC | Ncase = 135,458 vs.<br>Ncontrol = 344,901 |
| BD | Mullins et al.<sup>18</sup>,<br>2021 | PGC | Ncase = 41,917 vs.<br>Ncontrol = 371,549 |
| OCD | Arnold et al.<sup>19</sup>,<br>2018 | PGC | Ncase = 2,688 vs.<br>Ncontrol = 7,037 |
| PD | Forstner et al.<sup>20</sup>,<br>2021 | PGC | Ncase = 2,248 vs.<br>Ncontrol = 7,992 |

SPARK, Simons Foundation Powering Autism Research for Knowledge; PGC, Psychiatric Genomics Consortium;
ASD, autism spectrum disorder; ADHD, attention deficit/hyperactivity disorder; SCZ, schizophrenia; MDD, major depressive disorder; BD, bipolar disorder; OCD, obsessive-compulsive disorder; PD, panic disorder.

### Instrumental Variables Selection

The flowchart of the study is presented in Figure 1. Briefly, cord blood DNA methylation levels of *OR2L13,* which was associated with maternal GDM during pregnancy, were used as the exposure, and ASD, ADHD, SCZ, MDD, BIP, OCD and PD served as the outcomes respectively. To meet the core assumptions of MR analysis (i.e., i. Genetic instruments must be robustly associated with exposures of interest; ii. There were confounding factors of IVs and exposures. iii. IVs affected the outcomes only through exposures.), we performed the following data process and harmonization before MR analysis ^21^. Firstly, we obtained SNPs strongly associated with exposures (p < 1e-8). Secondly, the minor allele frequency (MAF) threshold of the variants of interest was 0.01 and palindrome SNPs were not included in our study. Thirdly, LD clumping process (r^2^ < 0.01 and clumping distance =10,000 kb) was conducted to ensure statistical independence between genetic variants. Finally, we identified 9 SNPs associated with cg20433858, 13 SNPs associated with cg03748376, 12 SNPs associated with cg00785941, 11 SNPs associated with cg04028570, 8 SNPs associated with cg08260406, 14 SNPs associated with cg08944170, and 11 SNPs associated with cg20507276.

**Figure 1.**
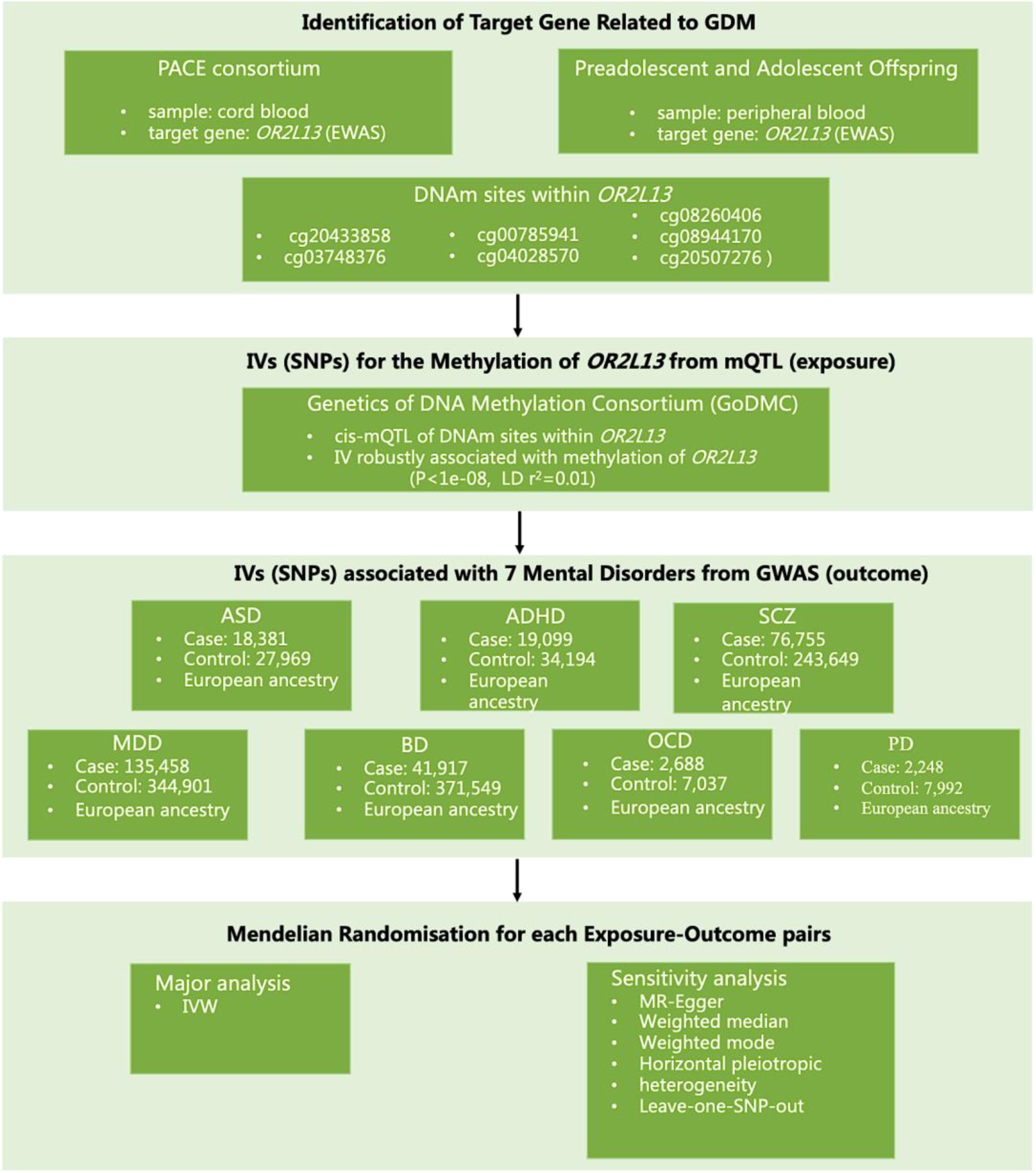
Study design overview for the two-sample MR analysis.

### Statistical analysis

The “TwoSampleMR” R package (version 0.5.6) was used to estimate the causal effect of cord blood DNA methylation on ASD, ADHD, SCZ, MDD, BIP, OCD and PD^22^. Inverse variance-weighted (IVW) method were applied as the main analysis methods. As our causal effect estimates might be affected by potential horizontal pleiotropic effects, we applied other MR methods (MR-Egger, weighted median, and weighted mode) as sensitivity analysis to assess the robustness of the MR main analysis results ^23–26^. Additionally, Cochran’s Q statistic was adopted in the IVW analysis to assess the potential heterogeneity between SNP-specific causal effect estimates related to blood DNA methylation ^27^. The intercept term in the MR-Egger regression was used to test for potential pleiotropy ^28^. Leave-one-SNP-out analysis was conducted to identify any single influential SNP that impacted the average causal effect estimate^29^.

### Ethical approval

This study is based on publicly available summary statistics from studies that had already obtained ethical approval; therefore, a separate ethical approval was not required.

## Results

Applying the fixed-effects IVW MR analysis, we found a significant causal association of lower *OR2L13* methylation with a higher risk of PD in offspring [cg03748376: odds ratio (OR)=0.81, 95% confidence interval (CI) =0.68-0.97, P =0.02; Figure 2]. However, for the remaining mental disorders, little evidence was found for a causal relationship between the methylation levels of *OR2L13* and these outcomes. (Figure 3, Supplementary Table S2).

**Figure 2.**
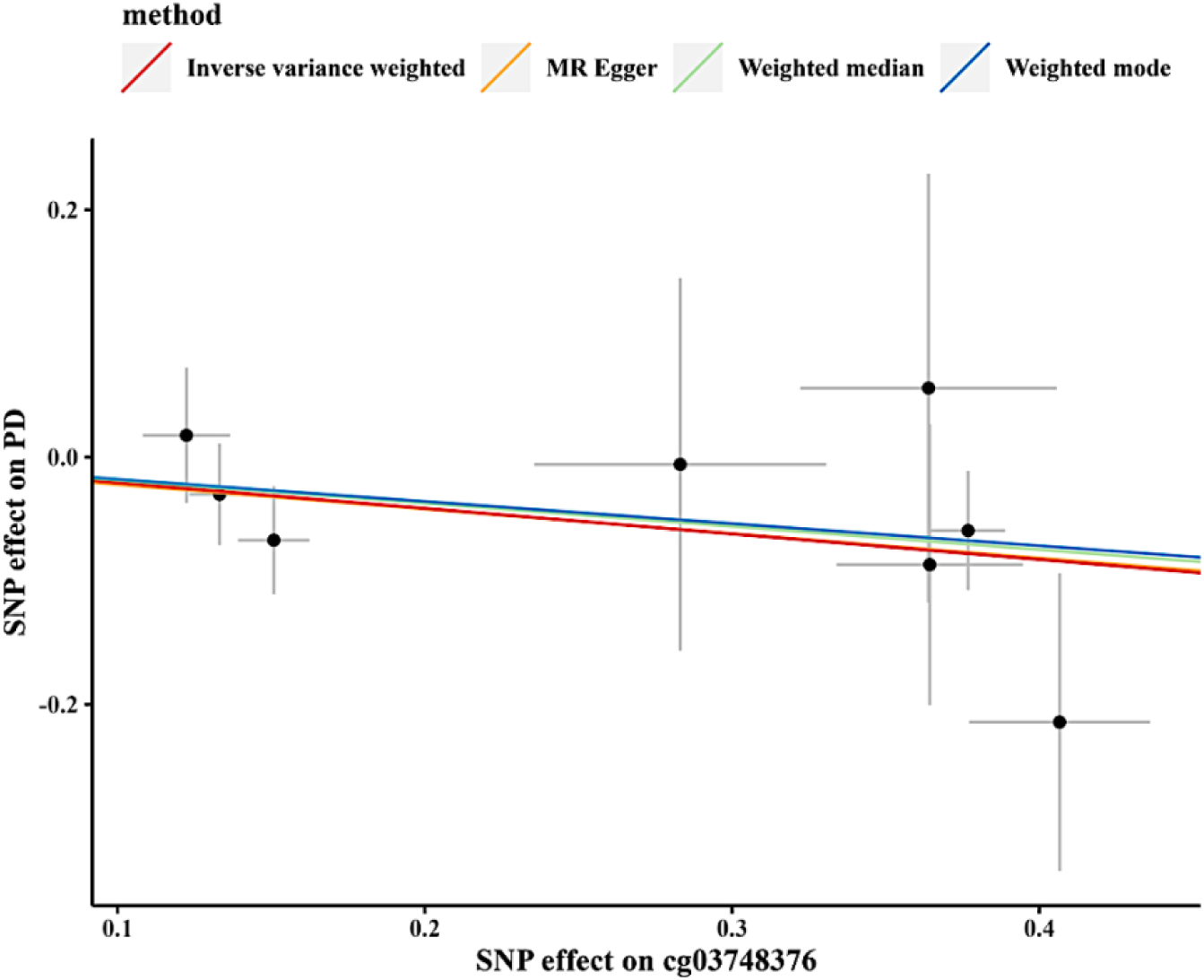
MR results of causal effects between *OR2L13* methylation (cg03748376) and panic disorder (PD). Scatter plot of genetic correlations of methylation level of cg03748376 and PD using different MR methods.

**Figure 3.**
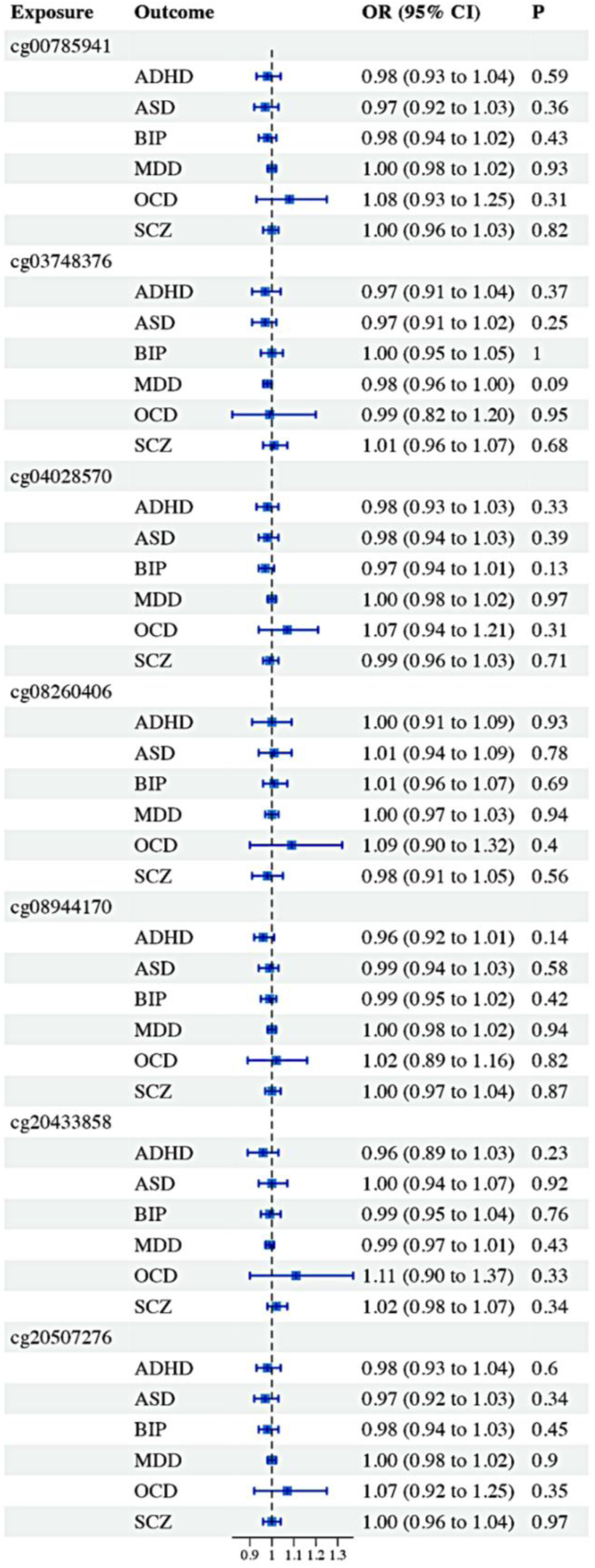
Two-sample Mendelian Randomization reveals little causal evidence for methylation of *OR2L13* on ASD, ADHD, SCZ, MDD, BIP and OCD.

Results of sensitivity analysis broadly support our main analysis results (Figure 2, Table 2). Cochrane Q statistics showed no evidence of heterogeneity for MR-Egger and IVW test in the causal relationship between *OR2L13* methylation (cg03748376) on risk of PD in offspring (p > 0.05) (Table 3). There was no horizontal pleiotropy in MR analysis results by using MR-Egger regression intercept approach (p > 0.05) (Table 3). The results of the leave-one-SNP-out analysis indicated that the IVW results were not driven by a single SNP (data not shown).

**Table 2.** Mendelian randomization (MR) results of causal effects between methylation of 7 targeted CpG sites and PD.

| **CpG sites**<br>**(exposure)** | **Methods** | **No. of**<br>**SNPs** | **OR** | **95% CI** | **P value** |
| --- | --- | --- | --- | --- | --- |
| cg00785941 | MR Egger | 10 | 0.78 | 0.49-1.24 | 0.32 |
|  | Weighted Median | 10 | 0.85 | 0.69-1.05 | 0.14 |
|  | IVW | 10 | 0.87 | 0.73-1.04 | 0.12 |
|  | Weighted Mode | 10 | 0.83 | 0.64-1.07 | 0.18 |
| cg03748376 | MR Egger | 8 | 0.82 | 0.56-1.21 | 0.36 |
|  | Weighted Median | 8 | 0.83 | 0.67-1.03 | 0.09 |
|  | **IVW** | **8** | **0.81** | **0.68-0.97** | **0.02** |
|  | Weighted Mode | 8 | 0.84 | 0.66-1.06 | 0.18 |
| cg04028570 | MR Egger | 10 | 0.87 | 0.63-1.20 | 0.42 |
|  | Weighted Median | 10 | 0.87 | 0.73-1.04 | 0.13 |
|  | IVW | 10 | 0.88 | 0.77-1.01 | 0.08 |
|  | Weighted Mode | 10 | 0.87 | 0.70-1.08 | 0.23 |
| cg08260406 | MR Egger | 6 | 0.78 | 0.32-1.89 | 0.61 |
|  | Weighted Median | 6 | 0.94 | 0.68-1.29 | 0.69 |
|  | IVW | 6 | 0.91 | 0.67-1.25 | 0.56 |
|  | Weighted Mode | 6 | 1.21 | 0.64-2.28 | 0.58 |
| cg08944170 | MR Egger | 13 | 0.95 | 0.63-1.43 | 0.81 |
|  | Weighted Median | 13 | 0.84 | 0.69-1.03 | 0.09 |
|  | IVW | 13 | 0.86 | 0.73-1.02 | 0.08 |
|  | Weighted Mode | 13 | 0.78 | 0.61-1.02 | 0.09 |
| cg20433858 | MR Egger | 6 | 0.81 | 0.59-1.10 | 0.25 |
|  | Weighted Median | 6 | 0.86 | 0.70-1.06 | 0.16 |
|  | IVW | 6 | 0.88 | 0.73-1.05 | 0.14 |
|  | Weighted Mode | 6 | 0.86 | 0.69-1.08 | 0.25 |

**Table 2 (continued). Mendelian randomization (MR) results of causal effects between methylation of 7 targeted CpG sites and PD.**
| **CpG sites**<br>**(exposure)** | **Method** | **No. of**<br>**SNPs** | **OR** | **95% CI** | **P value** |
| --- | --- | --- | --- | --- | --- |
| cg20433858 | MR Egger | 9 | 0.87 | 0.56-1.36 | 0.56 |
|  | Weighted median | 9 | 0.86 | 0.70-1.05 | 0.13 |
|  | IVW | 9 | 0.88 | 0.75-1.03 | 0.11 |
|  | Weighted mode | 9 | 0.84 | 0.67-1.06 | 0.18 |

**Table 3.**
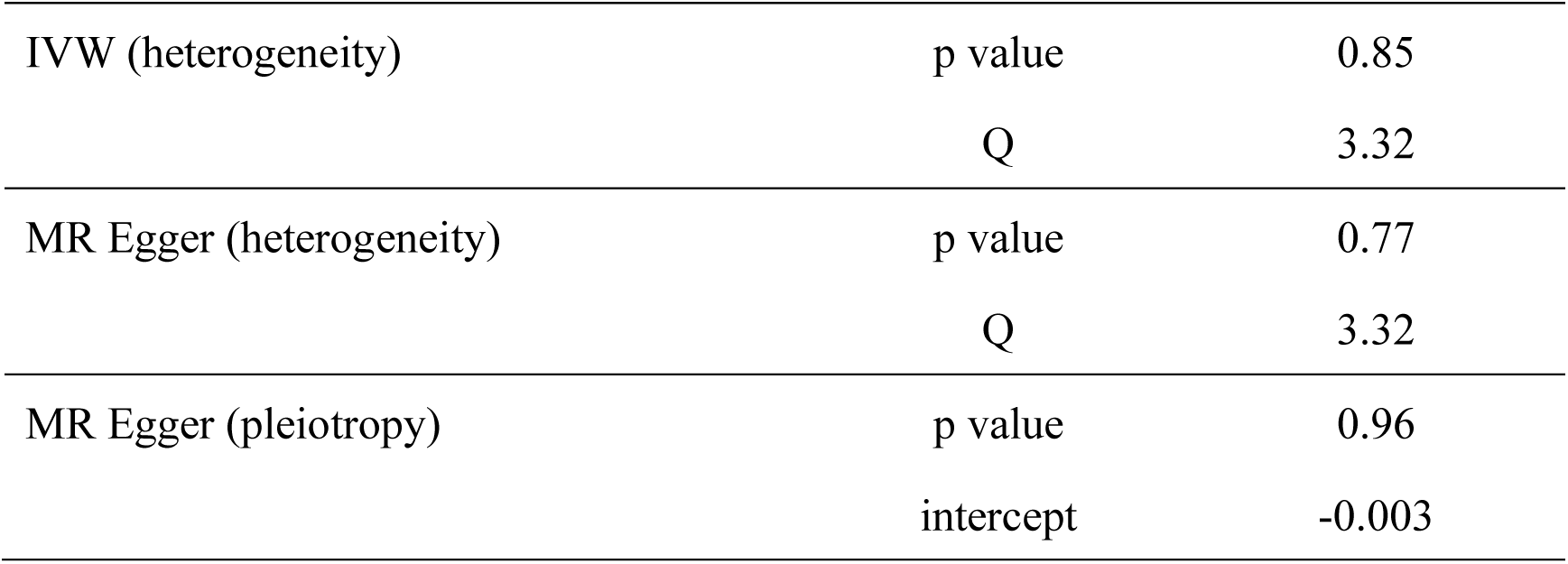
Sensitivity analysis of the Mendelian randomization (MR) analysis results of cg03748376 and PD.

|  |  |  |
| --- | --- | --- |
| IVW (heterogeneity) | p value | 0.85 |
|  | Q | 3.32 |
| MR Egger (heterogeneity) | p value | 0.77 |
|  | Q | 3.32 |
| MR Egger (pleiotropy) | p value | 0.96 |
|  | intercept | -0.003 |

## Discussion

To the best of our knowledge, this is the first MR analysis to investigate whether methylation used as proxy for GDM is causally associated with ASD and related mental disorders. Using genetic instruments from the largest available mQTL summary statistics of 32,851 participants, we found genetic liability to the lower *OR2L13* methylation serving as proxy for GDM causally associated with PD in offspring. This is consistent with previous population-based cohort studies, which have found that children born to mothers with diabetes during pregnancy were at increased risk of developing anxiety disorders^1, 30^. And two studies showed that GDM were associated with anxiety only in combination with maternal obesity, possibly triggering the inflammatory processes in the uterus and leading to damages to brain^31,32^.On the other hand, olfactory receptor 2L13 encoded by *OR2L13* in humans could interact with odorant molecules in the nose, initiating a neuronal response that triggers the perception of a smell ^33^. And experiments in vivo have showed that olfactory deficits cause anxiety-like behaviors in mice^34^.

However, we found little evidence to support a causal association of maternal GDM related *OR2L13* methylation levels change with other psychiatric disorders including ASD, ADHD and SCZ et al., which was inconsistent with previous association findings in conventional observational studies ^35–37^. It is possible that the previously observed positive association between GDM and these psychiatric disorders is coincidental or is confounded by unknown factors and not directly caused by GDM or methylation derived from GDM. Furthermore, while multiple studies have reported that methylation of several risk genes related mental disorders, including *BDNF*, *MAOA* and *SLC6A* and so on ^5^. Only *OR2L13* related to ASD was identified by Howe et al. using cord blood samples, which however, may not mirror methylation of different prenatal stages stems from hyperglycemia in utero. Deeper and more extensive research is needed to investigate these possible links.

## Limitations

This study has some limitations worth noting. First of all, due to the original data of cord blood related to GDM was not public, we used the indirect summary data of methylation as exposure instead. To avoid errors, further studies should use genetic markers of cord blood related to GDM. Additionally, the subjects included in this study are European descents, thus it should be cautious to generalize the findings in this study to population of other ancestry.

## Implications

Our study suggests that genetic predisposition to GDM is associated with the risk of PD in offspring. There was no evidence for a causal effect of GDM and a series of major psychiatric disorders (ASD, ADHD, SCZ, MDD, BD and OCD). And future studies are needed to clarify the putative causal mechanisms.

## Supplemental Materials

Data file S1. Supplemental tables (a single excel including 2 sheets as 2 Supplemental Tables).

Table S1: Single-nucleotide polymorphisms (SNPs) significantly associated with CpG sites of the *OR2L13* promoter (7 probes).

Table S2: Association between the methylation of *OR2L13* on ASD, ADHD, SCZ, MDD, OCD and BIP in offspring based on IVW method.

## Data availability

This study is based on publicly available summary statistics.

## Author contributions

FL and JZ conceptualized the study; XD, FL and JZ performed the analysis. FL, JZ, XD, LL, TR wrote the manuscript with contributions from all authors. All authors read and approved the final manuscript.

